# Hamdard Force: A scalable, data-informed community-based intervention to develop a mental health workforce in Pakistan

**DOI:** 10.1101/2025.07.17.25331689

**Authors:** Asma Humayun, Arooj Najmussaqib, Noor ul Ain Muneeb

## Abstract

**Background:** As part of a MHPSS initiative, this study presents the process of developing a scalable community-based mental health intervention - Hamdard Force – which involved training and monitoring the performance of a community workforce. The Psychological First Aid (PFA) has been adapted for Pakistan to develop the digital resources for this intervention. Our objective was to train the Hamdard Force to provide basic psychosocial support, identify people with mental healthcare needs, and refer them to relevant services.

**Methods:** We used a multi-method design guided by the ADAPT implementation science framework to systematically adapt PFA and develop a scalable community intervention in four stages. In the first stage, we identified the constraints and facilitators for PFA implementation. To address these challenges, we adapted PFA across three domains of structure, content, and digital design. In third stage, we tested the process of recruitment and training of Hamdard Force. Lastly, we designed and tested a Hamdard Force mobile application for seeking supervision, referring people to the MHPSS service, and seeking support for their well-being.

**Results:** The implementation challenges included gaps in mental health literacy; knowledge-to-practice skills; cultural and healthcare context; and for scalable and sustainable implementation. We used the FRAME approach to report the adaptations. For structural adaptation, we developed the guide into a modular design with interactive quizzes. We modified and simplified the content and added step-by-step instructions to convert it into a practical guide. Digital adaptation entailed converting the content into online courses in Urdu and English, incorporating audiovisual content and tools for course evaluation. We tested the intervention for recruitment, training and supervision of Hamdard Force with encouraging insights.

**Conclusion:** Hamdard Force offers a scalable, community-based intervention to strengthen mental healthcare. Through digital resources, the intervention demonstrates a mechanism for effective implementation in low-resource settings.

## Introduction

The World Health Organization calls for an active engagement of the community for integrated mental health programs to address the gap between the burden of mental disorders and access to appropriate mental healthcare (World Health Organization, 2021b). Even in high-income countries (HICs), access to mental health care is limited, where only a third of those with a common condition like depressive disorder receive treatment (Moitra et al., 2022). The challenge is far greater in low- and middle-income countries (LMICs) where the treatment gap for common mental disorders is estimated at 85% as compared to 40% in HICs (Ndetei et al., 2023). Like other LMICs, a major barrier for integrating mental health into primary healthcare is prioritization of scarce resources for tertiary healthcare (Hussain et al., 2018; Irfan, 2013; World Health Organization, 2018).

The objective of integrating mental health services in community settings in LMICs is to promote accessibility, acceptability, scalability of mental healthcare, and improve clinical outcomes, including treatment adherence (Goldberg & Huxley, 2012). The need to develop scalable community mental healthcare is even more critical for countries facing humanitarian challenges (World Health Organization, 2022), with Pakistan being one example (Riaz et al., 2023; UNHCR, 2023; World Bank, 2022). Effective implementation of community-based interventions, however, requires collaboration between state and donor agencies to scale up capacity building of health workforce and first-line responders through culturally adapted training resources (Philippe et al., 2022; World Health Organization, 2023).

Recommended by Inter-Agency Standing Committee (IASC), the Psychological First Aid (PFA) Guide is an evidence-informed, humane, and practical approach to provide basic psychosocial support in the community, usually as an early intervention following a humanitarian crisis (Movahed et al., 2023; World Health Organization, 2011a). Although widely used, the implementation, evaluation, and outcomes of most of the PFA programmes lack clarity (Wang et al., 2021), raising questions about its generalizability, standardization, and culturally competent curriculum (Ni et al., 2024).

The need to implement the PFA Guide as a community mental health intervention upsurged during the COVID-19 pandemic around the world, including Pakistan (Zafar et al., 2019; Chandler et al., 2023; Huque et al., 2021; Minihan et al., 2020). However, without contextual adaptation of PFA to local healthcare system, resources and cultural needs, a critical gap exists for its effective implementation (Sim & Wang, 2021).

In addition to the sociocultural dynamics, key barriers identified in LMICs include a lack of adequately trained personnel, effective coordination and communication mechanisms, health system preparedness, and political will (Anyebe et al., 2021; Li & Chen, 2022). Beyond the logistical and administrative constraints, insufficient stand-alone trainings without offering supervision (Wang et al., 2021) and gaps in implementing evidence-driven tools for monitoring and evaluation of PFA programmes have been reported (Castillo et al., 2019).

During the COVID-19 pandemic, the Ministry of Planning, Development & Special Initiatives (MoPD&SI) developed a comprehensive, evidence-based, and scalable model to provide mental health and psychosocial support (MHPSS) service in Pakistan (MoPDSI, 2022). As part of this initiative, a community intervention - *Hamdard Force,* was designed to train a mental health workforce. Hamdard is an adjective in Urdu word meaning “empathetic” and the title *Hamdard Force* describes a dual focus on the intervention and the workforce implementing it, deemed appropriate due to its cultural relevance and acceptance. The objectives of Hamdard Force were to provide basic psychosocial support, identify the mental health needs of people, and connect them to relevant services.

In this paper, we aim to present the process of developing the Hamdard Force as a community-based mental health intervention.

## Method

This study was conducted as part of the MHPSS project initiated by the MoPD&SI (letter no. 6(262) HPC/2020) and was led by AH. The objective of this study was to contextualize the PFA Guide to develop and test training resources to develop a scalable community-based intervention. We used a multi-method design informed by an implementation science framework - ADAPT guidance, a four-stage structure for systematically adapting interventions to new contexts (Moore et al., 2021).

### Stage 1: To assess the rationale for developing a training resource for a context-specific community intervention

We formed a core adaptation team of three psychiatrists and a clinical psychologist with expertise in community-based mental health interventions and contextual knowledge of local mental healthcare. They steered the adaptation process; coordinated multi-stakeholder consultations across the stages of ADAPT process; and finalized training resources. It adopted a consensus-based, co-production approach for all decision-making (Hawkins et al., 2017). The team performed a desk review, two Focus Group Discussions (FGDs), and four Key Informant Interviews (KIIs) to explore challenges and lessons learned from prior implementations of the PFA Guide in other contexts. The desk review spanned three databases (PubMed, Medline, and PsychINFO) and a manual review of grey literature from the humanitarian sector on implementing community-based mental health interventions.

All participants participated voluntarily in this study. The core team moderated the discussions to explore gaps in mental health literacy; needs for capacity building through training and supervision; and options to establish a referral system. For the FGDs, the participants were identified through professional networks based on their roles; interest in MHPSS, and availability (purposive sampling). For the KIIs, the participants were selected through snowball sampling. We used semi-structured questionnaires, and all sessions were audio-recorded with participants’ consent. A total of 26 participants were included in two FGDs (4 school teachers, 5 university students, 6 frontline healthcare workers, 4 MHPSS officers of humanitarian agencies, 4 people with lived experiences and 3 policy experts). KIIs were conducted with four experts (1 psychiatrist, 2 MHPSS officers and 1 health system manager). The core team reviewed transcriptions and analyzed the data to evaluate implementation constraints and facilitators using an iterative framework analysis (Gale et al., 2013).

Our study employed participatory research approach with multi-stakeholders, therefore, we used method triangulation to enhance validity of our findings. Thematic data saturation was monitored to guide the final sample sizes (Rahimi & khatooni, 2024). For thematic analysis, we employed Braun and Clarke’s (2006) six-phase framework using an inductive approach. Two members independently generated initial codes using Excel sheets, explored patterns, and organized them into themes through collaborative discussions with a third member. We ensured triangulation by comparing themes across FGDs and KIIs. Our analysis revealed five themes: Mental Health Literacy; Knowledge-to-Practice Gaps; Gaps in cultural context; Gaps in healthcare context; Sustainability and Scalability.

### Stage 2: To adapt the PFA Guide and develop training resources

The process of adaptation of the PFA Guide to develop our training resources is described below:

#### 2.1 Identify key adaptation considerations

Based on ADAPT checklist (Moore et al., 2021), the team formulated a semi-structured questionnaire to map considerations in our context and propose changes for adapting the structure and content of PFA (*see supplementary material A*).

#### 2.2 The adaptation process

An adaptation workshop was designed to review, synthesize and finalize the proposed adaptations using Delphi approach (Jorm, 2015). Through snowball sampling, a new group of experts were selected to ensure an independent perspective. These included 2 senior psychiatrists, 3 MHPSS field experts and 2 healthcare managers. Experts were briefed about the Hamdard Force initiative, identified gaps and the proposed changes. After consensus was obtained on proposed adaptations, the core team proceeded with the adaptation process across three domains: 1) Structural 2) Content and 3) Digital. Structural adaptation was required for clarity, improving organization and facilitating modular training. The content adaptation was conducted to address gaps in knowledge, knowledge to practice, cultural and healthcare contexts. The digital adaptation aimed to convert the adapted guide into online courses via the Moodle platform to address the need for a scalable modality of training (Details in **Table 1**).

**Table 1.** Adaptation of PFA to develop HF courses.

| **1 STRUCTURAL ADAPTATION** |  |  |  |
| --- | --- | --- | --- |
|  | **Rationale** | **Original content in PFA (Chapter)** | **Adaptations in HF course (Module)** |
| 1.1 | Modular restructuring for optimized utility by organizing content into focused units, each aligned with specific learning objectives. | Five chapters:<br>1. Understanding PFA;<br>2. How to help responsibly;<br>3. Providing PFA;<br>4. Caring for yourself & your colleagues;<br>5. Practice what you have learned. | 10 modules excluding Introduction:<br>1. Hamdard force: Code of conduct;<br>2. About your training;<br>3. Look;<br>4. Listen;<br>5. Link;<br>6. Techniques to help;<br>7. Children and adolescents;<br>8. People with mental health conditions & disability;<br>9. People at risk of discrimination or violence;<br>10. Care for yourself. |
| 1.2 | The principles of PFA are converted into a module each by integrating clear *How to do* instructions. | Providing PFA – look, listen and link (3) | Developed into Modules 3 - 5. |
| 1.3 | The guidelines to help vulnerable population were covered into three separate modules. | People who likely need special attention (3) include children and adolescents, people with health conditions or disabilities and people at risk of discrimination or violence | Separate modules (7-9) added to provide guidelines to support children and adolescents; people with mental health conditions & disability; & those at risk of discrimination or violence. |
| 1.4 | Within a module, the content was restructured to ensure clarity and promote comprehension. | Providing PFA (3)<br>Covered in one section | Techniques to help (6)<br>Following sections incorporated with clear instructions to communicate: |
|  |  |  | -Help people think clearly to solve their problems<br>-Strengthen their support system<br>-Help cope with stress |
| 1.5 | To strengthen learning experience; promote knowledge retention; reinforce concepts; and encourage critical thinking. | Practice what you have learned (5)<br>Case scenarios | Instead of case scenarios (more suitable for in-person discussions), 15 interactive quizzes added to assess knowledge retention within each module. |
| 1.6 | To develop a scalable intervention, the duration of the course was optimized. | - | Theoretical content summarized by incorporating concrete instructions without compromising the core content. |
| **2** | **CONTENT ADAPTATION** |  |  |
| **2.1** | **Gaps in mental health literacy** |  |  |
| 2.1.1 | Changes in behaviour enlisted to help identify people with mental health needs. | People who likely need special attention (3)<br>Section on people with health conditions & disability included | **People with mental health conditions & disability (8)**<br>*Who should be referred, e.g. people who are so distressed that they are unable to function normally and care for themselves; not sleeping well; confused and disorientated; restless and agitated; threatening to harm themselves or others; using excessive amounts of harmful substances including alcohol; not taking their regular medication; experiencing violence or are being abused.* |
| 2.1.2 | Content added to explain at risk population, impact of violence and instructions to help people at risk. | People who likely need special attention (3) | **People at risk of discrimination or violence (9)**<br>-Common examples of discrimination or violence<br>-Impact of violence<br>-Detailed instruction for helping them e.g., helping them meet their basic needs – food, safe housing, aid, ration, safety and connecting them to health facility, |
|  | Exposure to violence and trauma is common in our humanitarian context.<sup>1</sup> |  | family or trusted people, police, legal help, or to MHPSS service. |
| 2.1.3 | Content was expanded **to clarify concepts related to awareness and stigma** | Caring for yourself and your colleagues (4)<br>Includes a general paragraph | Sections were added to Care for yourself (10)<br>*Why is it important to look after yourself?*<br>*What are the common sources of stress?*<br>*Which steps should be taken when you feel unwell or exhausted?* |
| **2.2** | **Knowledge to practice gaps**<br>**Prompts were added using simple language to improve learner engagement & clarity; and strengthen communication skills.** |  |  |
| **2.2.1** | Example 1<br>**To strengthen communication skills and protect the rights of the person** | Providing PFA Listen (3)<br>General guidelines | Listen (4)<br>The following prompt was added<sup>2</sup>:<br>*Introduce yourself: My name is ----- and I am working as a volunteer with ----- to help link people with sources of help.*<br>*Are you willing to speak with me? Our conversation can last for around 30 minutes.* |
| **2.2.2** | Example 2<br>**To strengthen communication skills** | Providing PFA - Ask about people's needs and concerns (3)<br>General guidelines | Identify their needs and concerns (4)<br>*How are you feeling? Please describe your problems.*<br>*Is there anything else that worries you? Are there any practical concerns that worry you?* |

<sup>1</sup> (Humayun et al., 2025).
<sup>2</sup> (World Health Organization, 2018)

|  |  |  |  |
| --- | --- | --- | --- |
| **2.2.3** | **Example 3**<br>**To address gaps in psychological literacy.** | Providing PFA (3)<br>General guidelines to help people cope with problems:<br>*A person in distress can feel overwhelmed with worries and fears. Help them to consider their most urgent needs, and how to prioritize and address them.* | Techniques to help (6)<br>Instructions to inquire about the problem added.<sup>3</sup><br>*a) What problems are you facing at the moment?*<br>*b) Which problem would you like to work on first?*<br>*c) What are the possible solutions to the problem?*<br>*d) Have you solved similar problems in the past? If so, how did you do it?* |
| 2.2.4 | Instructions were added to strengthen skills to help adolescents during a crisis. | Providing PFA - Children, including adolescents (3)<br>General guidelines | An example for Children and Adolescents (7)<br>If a 15-year-old shares their fear – What if someone in my family becomes unwell? You should respond by saying:<br>*“Yes, it is possible that someone in your family may become unwell.*<br>*Let’s discuss what we can do to protect ourselves.*<br>*Let’s discuss how other people we know dealt with such a situation”.* |
| **2.3** | **Gaps in cultural context** |  |  |
| **2.3.1** | The entire guide is adapted considering the healthcare and cultural context to enhance conceptual understanding. | Understanding PFA (1) | -Relevant options for in-person and remote settings to provide PFA enlisted (2)<br>-Psychosocial reactions in cultural context added e.g., People may not be able to grieve by not being able to organize burials, engage in funerals etc. |

<sup>3</sup> World Health Organization. (2018). *Problem management plus (PM+): individual psychological help for adults impaired by distress in communities exposed to adversity. (Generic field-trial version 1.1).* https://www.who.int/publications/i/item/problem-management-plus-(-pm-)-individual-psychological-help-for-adults-impaired-by-distress-in-communities-exposed-to-adversity

|  |  |  |  |
| --- | --- | --- | --- |
| 2.3.2 | To protect rights of people in cultural context.<br>To respect social norms<br>To respect beliefs and religion<br>To protect the rights of vulnerable groups in the cultural context | How to help responsibly - Respect safety, dignity and rights (2) | Some examples (1):<br>*-You should not accept money or favors in return for your help (1).*<br>*-You should not impose your beliefs or religious interpretations of the crisis on people.*<br>*-If you help organize a burial, respect the deceased and provide support to the surviving family members.*<br>*-Facilitate people to connect with spiritual leaders or perform spiritual rituals (5)*<br>-Added examples in marginalized groups like religious minorities, transgender community, women facing domestic violence (9) |
| **2.3.3** | In view of limited mental health literacy, specific responsibilities were not considered relevant. | Providing PFA - Help people to feel calm (3) | Removed techniques for breathing and mindfulness. These interventions are covered under supervision of clinical psychologists in our service (See 2.4.2) |
| **2.4** | **Gaps in healthcare context** |  |  |
| 2.4.1 | Theoretical content about PFA was adapted into competency-based learning outcomes to ensure practical skills development. | Understanding PFA (1)<br>Theoretical description of PFA | Competency-based learning outcomes added in 7 bullet points to explain the core skills that participants will be equipped after training e.g., skills to provide basic support, and practical support to people affected by crisis (2). |
| 2.4.2 | To support HF members & monitor their baseline mental health. | Caring for yourself & your colleagues - Managing stress (4) | Instructions to use MyCare+ app for stress management.<sup>4</sup> (10)<br>A questionnaire (GHQ-12) added.<sup>5</sup> |

<sup>4</sup> Humayun et al., 2020. https://journals.lww.com/invn/fulltext/2020/18020/designing_psychosocial_support_for_covid_19.7.aspx
<sup>5</sup> (Goldberg & Williams, 1988). https://www.scirp.org/reference/ReferencesPapers?ReferenceID=189650

|  |  |  |  |
| --- | --- | --- | --- |
| 2.4.3 | To address implementation gaps, mechanisms to standardize and regulate service providers incorporated. | - | Added:<br>Inclusion criteria and steps to join the HF; directory for support services; roles & responsibilities of service providers. |
| 3 | **Digital adaptation** |  |  |
|  | **Feature** | **Rationale** | **Digital adaptation** |
| 3.1 | Self-enrollment online course | To develop a scalable and cost-effective training course which offers flexible access to a wider target and ensures consistent learning opportunity accommodating diverse schedules. | The adapted guide was converted into an online course (Moodle) using slide format. |
| 3.2 | Language | Limited proficiency in the English language of the target audience | Another course was developed into Urdu language. |
| 3.3 | Duration of the course | Ensure feasibility to engage **community workers and reduce dropout rate making it suitable for emergency settings.** | The courses were designed to be completed within 2 hours.<sup>6</sup> |
| 3.4 | Pedagogy | Interactive pedagogy to ensure comprehension for audience with low-literacy rate and attention.<br>A deliberately simple, approachable visual language designed to communicate care, | Audio-visual content incorporated, illustrations were added in the course, along with audio recordings.<br>Illustrations were created using a signature SS-style (specific visual style) doodle. |

<sup>6</sup> During the feasibility testing in ICT, the course completion report showed that some participants completed the course over more than one session (taking more than 2 hours).

|  |  |  |  |
| --- | --- | --- | --- |
|  |  | connection, and accessibility for people with ill mental health. |  |
| 3.5 | Interactive activities/ quizzes | Integrated activities/ quizzes for reinforcing knowledge retention and encouraging critical thinking. | Assessment quizzes (MCQs/ Fill in the blank/ other types of questions) and activities (labeling, matching etc.) were configured in all modules in a creative format. |
| 3.6 | Assessment questionnaire | Pre- and post-training knowledge assessment to evaluate the effectiveness of the course. | The adaptation is described in the next section of results. |
| 3.7 | Feedback report | To establish acceptability, effectiveness and fidelity of the assessment. | Feedback form on a three-point Likert scale was added to assess the following: course design, content, language, duration, quizzes, audio recordings and illustrations.<br>The form also invites qualitative feedback through an open-ended prompt. |
| 3.8 | Course Report (Learners with their status) | Insight about the demographic details of the trainees as well as their status, duration and extent of course completion. | Report containing ID, name and email address with completion status of entire course or module, and duration of course completion. |
| 3.9 | Certificate | Incentivize the participants to complete the course and seek supervision for at least 10 cases and recognize their contribution towards community intervention. | Access to download E-certificate after completing training requirements. |

#### 2.3 The translation process

The adapted guide was translated into Urdu using WHO guidelines (World Health Organization, 2020). A bilingual team, comprising a psychiatrist and a psychologist, translated the original English draft (M1) individually and then synthesized into a cohesive Urdu version (M2). Subsequently, another team of two bilingual mental health experts performed the back-translation of M2 into English for comparison with the original (M1). Through this direct comparison, the team reviewed discrepancies in grammar, meaning, and language usage, thus streamlining the translation process.

### Stage 3: To evaluate the training resource and test the feasibility of the intervention

The Islamabad Capital Territory (ICT) was identified as the pilot site for feasibility testing and followed these steps:

#### 3.1 Recruitment in Hamdard Force

We performed a structured exercise using a stakeholder mapping matrix, informed by a review of official websites of relevant public and private sector organizations. This was followed by a stakeholders’ meeting to discuss objectives of recruitment and training. The partner organizations helped to identify relevant service providers and develop a service directory for ICT, which was integrated into the Hamdard Force courses. The directory provided immediate access to necessary support services and includes contact information for emergency support, counseling, shelter care for vulnerable population, financial aid, and legal services. They also contributed to form an inclusion criterion for recruiting Hamdard Force (*see supplementary material B*). Following the meeting, an instructional video of the recruitment and training process was circulated, and eligible participants registered through a form on the web-portal. A Hamdard Force contact directory was also formed to send bulk emails/SMS notifications to the registered members.

#### 3.2 Training of Hamdard Force

Once registered, participants accessed and enrolled in the Hamdard courses on the Learning Management System (LMS). The training effectiveness was evaluated using both pre-post assessments of knowledge and feedback from the participants. For this purpose, a 20 items knowledge questionnaire was developed. This included 10 true/false statements from the PFA Facilitators’ Guide (2013) and 10 MCQs were designed from the adapted guide to increase reliability of assessment and reduce guessing probability. The questionnaire demonstrated acceptable reliability (r=0.70), and a paired sample t-test assessed pre-post knowledge scores. Both qualitative and quantitative (seven items rated on a three-point Likert scale) feedback from the participants was collected via an online survey form.

### Stage 4: To implement and maintain the intervention at scale

To ensure scalability and cost-effectiveness, our digital components included a database of registered Hamdard Force and online training courses with monitoring of outcome indicators. Another vital component for scaling up our intervention was the Hamdard Force mobile application. Upon completion of their course, participants received a link to download the application, to refer cases in real time to the MHPSS web-portal; seek supervision; provide feedback; and access the guide and the service directory. During the feasibility testing, they were required to refer and seek supervision on at least 10 cases to download their training e-certificate from the LMS.

## Results

### Stage 1: To assess the rationale for developing a training resource for a context-specific community intervention

Based on the outcomes of desk review, FGDs, KIIs, we present our analysis of the constraints and facilitators under five themes:

1) Gaps in mental health literacy: Participants reported limited understanding of community members about mental health problems, their impact, needs and rights of vulnerable populations, and available services. Negative attitudes and stigma were reported to be prevalent. However, individuals under 40 years who had greater exposure to information through digital platforms were reported to be more open to discuss mental health problems.
2) Knowledge to practice gaps: Some parts of the PFA Guide were discussed with the participants who understood ‘what needs to be done’ but found it difficult to apply psychosocial concepts e.g., there were ambiguities on how to ‘provide support’. They highlighted the need for clear, practical, stepwise guidance on ‘how to provide support’. Some participants already had basic skills through their ongoing work with vulnerable populations. It was pointed out that techniques like breathing and mindfulness might not be suitable for training. Despite these limitations, participants shared strong willingness and motivation to be trained and to provide help.
3) Gaps in cultural context: Participants highlighted cultural perspectives in relation to the PFA Guide. Participants reported that people may often be judgmental towards individuals with mental health crisis and impose their own religious beliefs, which discourages help-seeking. They suggested that the emotional reactions surrounding grief or culturally sensitive mourning practices should be incorporated in the training. Despite being aware of the discrimination faced by vulnerable groups, including religious minorities, transgender persons, and women facing domestic violence, their knowledge about their rights was limited. Some were even reluctant to engage with the marginalized population due to existing cultural and religious taboos. However, they acknowledged the need to address these issues through training.
4) Gaps in healthcare context: Lack of availability of contextualized training material, trainers, health system infrastructure, and financial resources were highlighted as key challenges. The groups suggested that existing community networks such as local influencers, teachers, college students and NGO field staff should be engaged. Some participants had already attended one-off trainings in the PFA Guide but felt that these were mostly didactic and ineffective in their skills development. They were unaware of any mechanisms for training evaluation for training efficacy or evidence-informed decision making in their practices.
5) Sustainability and scalability: Participants expressed concern that existing programs are not just very limited but operate in silos with absence of supervision, follow-up, and structured referral pathways. They were concerned about lack of funding to sustain such initiatives, and strategic planning for scale-up. They also emphasized the need to recruit diverse groups, particularly youth who are tech-savvy, for successful implementation of such intervention.

### Stage 2: To adapt the PFA Guide and develop training resource

We used the Framework for Reporting Adaptations and Modifications-Enhanced (FRAME) guidance to document and report adaptations to strengthen transparency and replicability *(see supplementary material C)*. The adaptation of PFA using this approach is presented in Table 1.

### Stage 3: To evaluate the training resource and test the feasibility of the intervention

The feasibility testing of the training resources and intervention was conducted in ICT through intersectoral collaboration. Our team conducted multiple in-person introductory sessions with lady health workers (LHWs), school teachers, students and other Hamdard Force partners to introduce the MHPSS initiative and discuss their role; and guide them on the process of registration, training and supervision. Our governmental partners nominated frontline healthcare workers, rescue staff, teachers, university students, and social service workers. We also received nominations from non-governmental and civil society organizations working with vulnerable and marginalized population including children living in urban slums, people with disabilities, victims of gender-based violence, and volunteers from rural councils.

During the testing period of three weeks, a directory of frontline workers was prepared (2100 nominations), and bulk SMS invitations with the link for registration were sent. Out of which, a total of 758 participants registered as Hamdard Force (**Table 2**).

**Table 2:** Demographic details of registered Hamdard force members (n=758)

| **Table 2: Demographic details of registered Hamdard force members (n=758)** |  |  |
| --- | --- | --- |
|  | **F** | **%** |
| **Age** |  |  |
| Under 18 | 182 | 24 |
| 18 – 29 years | 151 | 19.9 |
| 30 – 39 years | 130 | 17.2 |
| 40 – 49 years | 148 | 19.5 |
| 50+ years | 147 | 19.4 |
| **Gender** |  |  |
| Male | 397 | 52.4 |
| Female | 361 | 47.6 |
| **Profession** |  |  |
| Teacher | 215 | 28.4 |
| Healthcare | 125 | 16.5 |
| Mental Health professional (Counselors, Psychologists, etc.) | 28 | 3.7 |
| Student | 226 | 29.8 |
| Community Member | 41 | 5.4 |
| Housewife | 8 | 1.1 |
| Social Worker | 4 | 0.4 |
| Others | 107 | 14.1 |
| Unemployed | 4 | 0.4 |

Due to logistical difficulties, we only had two weeks to pilot the training resources. During this period, a total of 265 participants enrolled in the courses, 107 completed the courses and 208 submitted their feedback (*see supplementary material-D)*. An attrition rate of 22.6% was observed between pre-post training stages. The knowledge evaluation showed a significant improvement in post-test scores. The mean score increased from 11.49 to 12.15 (t = 3.66; p < .001).

The quantitative feedback supported the program’s overall design, content, and delivery methods (Figure 2). In qualitative feedback, participants offered suggestions such as improving the audio-visual components and refining the course design, particularly by adding a progress bar for each module. About 20% of users had varying views on course duration as either longer or shorter. Other recommendations included adding explanatory notes for correct quiz answers and offering revision sessions.

**Figure 2:**
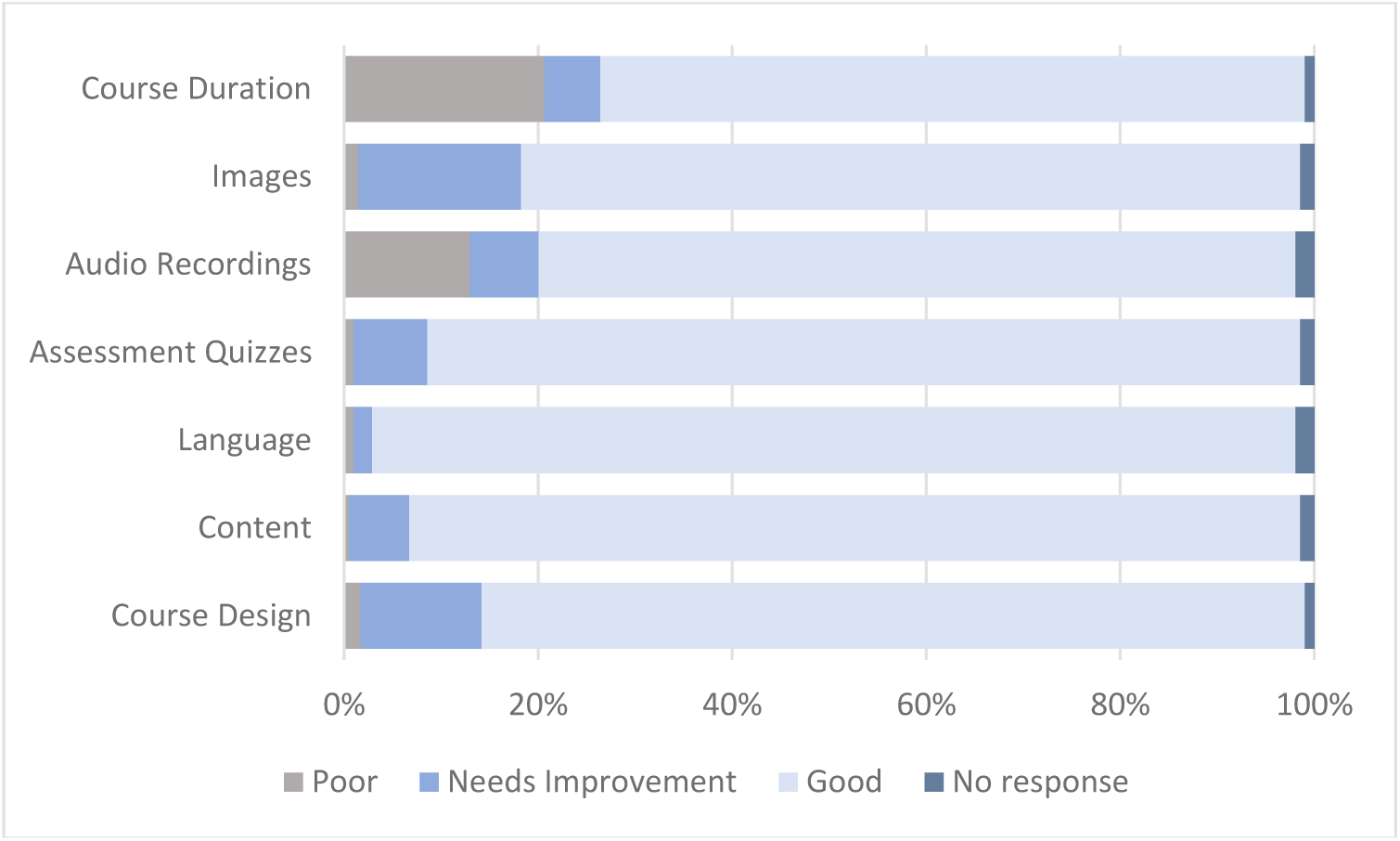
Quantitative feedback of the Hamdard Force courses

### Stage 4: To implement and maintain the intervention at scale

The Hamdard Force mobile application is a lightweight, low-bandwidth solution for use in resource limited settings. It supports continued functionality without internet access, which allows data to be securely stored and synchronized once the internet is available. The application provides bilingual (English and Urdu) support and is fully integrated with the centralized MHPSS web-portal. To refer cases, Hamdard Force members are required to seek permission from the people with mental health needs and send their demographic and contact details, along with a brief description of the problem or reason for referral. They are also required to complete General Health Questionnaire-12 (D. Goldberg & Williams, 1988) for the person who needs help to assess baseline mental health. During the two-week of testing training resources, 17 participants submitted 30 cases through the app, and requested supervision for 20 of these cases.

## Discussion

This qualitative study describes the process, challenges and lessons learned from developing a scalable community-based mental health intervention for resource-constrained settings.

The Hamdard force resources comprising a printed guide, online courses, and a mobile application, have been systematically adapted to align with the needs of the target audience, ensuring engagement for effective implementation (Giebel et al., 2024; IFRC, 2018; Wang et al., 2025). The adapted content and context-sensitive guidance is suitable for integrating our community intervention into routine healthcare practice (Berliner & Kolko, 2016; Wang et al., 2025). Furthermore, this intervention has the potential to include other targeted interventions and address the existing gaps in mental health support and the dearth of professionals in crises (Jamil & Khan, 2025; Kang & Choi, 2021; Yousuf et al., 2023). For instance, in health emergencies such as disastrous floods, training materials can be incorporated and deployed through this community-based intervention. The integration of digital resources with a centralized MHPSS portal provides a database to monitor and evaluate the performance of Hamdard Force while addressing critical data gap for informed policy-making (Dayani et al., 2024).

Following Giebel et al. (2024), we explored the constraints and facilitators for implementing a community-based mental health intervention by engaging multiple stakeholders for inclusive, comprehensive outcomes, and community ownership. We also agree with them that utilizing existing community workforce is crucial for smooth and sustainable implementation. Therefore, we developed a registration and data management mechanism that can even enable emergency deployment of trained workforce, when needed. We also defined a recruitment criteria suitable for varying levels of educational backgrounds and digital literacy skills among community members (Adewunmi et al., 2025; Khan et al., 2024).

Despite being widely endorsed compared to other available guides (Shultz & Forbes, 2014), the adaptation of the PFA Guide to local context is strongly recommended (World Health Organization, 2013). While the effectiveness and feasibility of implementing the PFA Guide for community workers is well established (Chandler et al., 2023; Shah et al., 2020; Wang et al., 2021), evidence on its adaptation and fidelity of implementation remains limited (Wang, Norman, Edleston, et al., 2024).

To overcome the gap in contextual adaptation of PFA, we systematically deliberated and documented each change and the supporting rationale using an implementation science framework (Moore et al., 2021; Wiltsey Stirman et al., 2019). The framework approach has been increasingly used to adapt, document and scale up evidence-based psychological interventions (Grubin et al., 2022; Sangraula et al., 2021; Wang et al., 2025), and to avoid unintended modifications that might remove the core content of interventions or misalign with the target population (McCrabb et al., 2019).

We restructured the PFA Guide to address the needs of our target audience (Grubin et al., 2022; IFRC, 2018; Wang et al., 2025). The adapted guide is presented in a modular format, dividing each module into clearly defined sections, highlighting actionable steps and incorporating bullet points for clarity. This structure helps the workforce navigate the content and apply knowledge effectively in implementation.

One of our challenges was to enhance conceptual understanding and cultural relevance while integrating detailed guidance on implementing stated actions, as content to build training skills is rarely designed or included in PFA training, hampering its effectiveness and quality (Horn et al., 2019).

The content adaptation addressed the need for practical guidance and sensitivity to cultural, religious, and social practices (World Health Organization, 2011b). Focusing on vulnerable populations, we also addressed risk factors such as grief, exposure to violence, and discrimination (Humayun et al., 2025). We incorporated step-by-step concrete instructions to address gaps in mental health literacy, and strengthen the skills of Hamdard Force to achieve learning outcomes (Humayun et al., 2025; Wang, Norman, Xiao, et al., 2024; World Health Organization, 2021a). Though the translation of the PFA in Urdu is available by the WHO, it is a literal word-for-word translation. Our experts highlighted its complex language and practical challenges in implementation for the target audience, who have limited literacy skills and more familiar with conversational Urdu.

Our preference for digital adaptation of the PFA Guide represents a paradigm shift from traditional face-to-face trainings across the country to overcome system inadequacies and resource limitations, significant barriers for implementing a scalable intervention involving in-person training and supervision (Giebel et al., 2024). Given the lack of dependable and accessible healthcare infrastructure coupled with growing digital access, a digital platform appeared to be the most viable solution for a scalable intervention (Philippe et al., 2022).

Though many training courses based on PFA are available online (IFRC, 2024; John Hopkins University, 2025), we recognized the need for a customized course optimized for content and duration to target indigenous cultural needs. Our course is available in bilingual format and integrated with a comprehensive digital MHPSS service. The audio-visual features were carefully designed, engaging mental health professionals and people with lived experiences, enhancing relevance and acceptability.

There is a consensus that PFA training without supervision leads to gaps in essential competencies (Wang et al., 2021). We intended to develop a cost-effective and sustainable mechanism (Buntrock, 2024; Fu et al., 2020) where a trained workforce could be supervised for making referral decisions in real time. For this reason, the low-bandwidth design and offline functionality of our mobile application promote accessibility for users.

A high global public health priority about engaging community workers in delivering mental health interventions is their well-being (Søvold et al., 2021), which gained much attention during the pandemic when substantial evidence came to light about the mental health risks of the front-line responders (Maple et al., 2024). Many countries resorted to digital solutions for providing individual and peer support to healthcare workers (Chandler et al., 2023; Drissi et al., 2021). We offer an additional feature in the mobile application to record the baseline well-being through a self-reported questionnaire (D. Goldberg & Williams, 1988), and offer help through a hybrid approach using another mobile application integrated with MHPSS system (Humayun et al., 2020). Unlike self-help chatbot services in humanitarian contexts that lack empirical evidence, clinical accountability, and transparency (Armitage, 2022), our intervention offers evidence-based support to frontline workers while providing supervised care and maintaining data privacy.

Our findings showed a significant improvement in knowledge post-training, which has also been reported previously (Lalani & Drolet, 2020; Movahed et al., 2023; Sijbrandij et al., 2020; Ting et al., 2024). The feedback from the participants reinforced the need for community participation, role of PFA training to address gaps in mental health literacy and stigmatizing attitudes, and willingness to be trained in supporting vulnerable communities, even beyond the humanitarian context.

High attrition rate is observed in other digital first aid studies, like in Ukraine (Asanov et al., 2024) and can be overcome by incentivizing participation. In our case, the attrition rate may be attributed to lack of motivation, challenges in digital accessibility, and financial constraints (Abbey et al., 2014). We are mindful of the attrition bias, and need of additional support for digital navigation for our target population, for instance, LHWs. Our online courses allow the participants to seek technical support during course completion to overcome digital exclusion, a common challenge observed in digital mental health solutions (Al Dweik et al., 2024). Nevertheless, the feedback indicated fidelity of training resources, which reinforces the need to scale up data collection and conduct more comprehensive evaluations. Following the completion of feasibility testing, we incorporated suggestions and revised training resources to enhance their applicability and functionality for future implementation.

### Limitations

Our training evaluation was limited to knowledge assessment. Since this alone cannot measure the PFA efficacy, future implementations should adopt stronger methodological designs such as Randomized Control Trials. They may also include additional outcome measures, such as confidence, attitudes, communication skills, and mental health indicators (Hermosilla et al., 2023; Wang et al., 2021).

Due to time and resource constraints, we only tested the online training courses. Whereas, disseminating the printed guides might help improve engagement and strengthen knowledge retention (Wang et al., 2021). Comparisons between self-directed online training and facilitated in-person training can provide better insights about effectiveness of training modalities and their cost-implications. For self-directed online courses, a pre-training webinar might have helped participants’ preparedness.

We partially tested the Hamdard Force service, whereas the implementation of the entire mechanism can provide more in-depth information on the efficacy of the intervention. Furthermore, in-depth cultural adaptation of the community intervention by incorporating local concepts of distress and help-seeking pathways may enhance its effectiveness and acceptability (Heim & Kohrt, 2019).

### Future directions

The ratio of trained community workers to the general population depends on the objectives of the intervention and community needs. In a populous country like Pakistan, an average district exceeds one million residents, training even 0.1% of the population means developing a workforce of over a thousand people. Given the scale of mental health needs, there is a dire need for a scalable and sustainable intervention.

Our efforts to fully test the intervention were partially successful due to funding constraints, which is a perpetual challenge in resource-constrained settings. Sustained scale-up requires strong policy commitment, particularly toward digital health infrastructure, integration of Hamdard Force intervention into routine healthcare, and national disaster plans.

We also need to strengthen our evaluation strategies for more effective implementation. Future studies may assess economic evaluation and long-term mental health outcomes in communities served by Hamdard Force.

In our case, the incentives offered for Hamdard Force included training certification, supervision, and support for their mental well-being. Opportunities for career development and monetary incentives will further help to sustain such community services.

Lastly, establishing a platform for multi-stakeholder collaboration is crucial for scale-up. We disseminated regular updates, but quarterly stakeholder reviews for strengthening partnerships and aligning long-term goals are important. As an outcome of these collaborations, pilot implementation of the Hamdard Force intervention is now being planned in additional provinces.

## Data Availability

The data that support the findings of this study are available from the Health Section at the Ministry of Planning, Development & Special Initiatives, Government of Pakistan. Confidentiality restrictions apply to the availability of the data used under the license for the current study, and so is not publicly available. However, the data can be made available from the authors upon reasonable request after formal approval from the Ministry of Planning, Development & Special Initiatives, Pakistan.

## Acknowledgments

We acknowledge major contributions towards the adaptation of the PFA Guide by Israr ul Haq, Faisal Rashid, and Sarah Rathore, who were part of the core adaptation team.

We would also like to thank Tuba Rahna, who was part of the initial thematic analysis.

This work was completed and tested as part of the MHPSS initiative supported by Dr M Asif, Chief Health, MoPD&SI, and funded by UNICEF in 2021.

We would also like to thank Ayesha Chaudhary, Director TechHive, and her team for their help in developing the digital resources.

## Ethics approval and consent to participate

This study was conducted as part of the Mental Health and Psychosocial Support Project, approved by the Ministry of Planning, Development & Special Initiatives in compliance with ethical standards and consent protocols under letter no. 6(262) HPC/2020.

## Consent for publication

All authors consented to publication.

## Authors

AH: Conceptualization, Methodology, Visualization, Supervision, Project administration, Writing –original draft, Writing – review and editing

AN: Methodology, Analysis, writing – original draft

NM: Data curation, writing – original draft

## Abbreviations

PFA: Psychological First Aid;
HICs: High-income Countries;
LMICs: low- and middle-income countries;
MoPD&SI: Ministry of Planning, Development & Special Initiatives;
MHPSS: Mental Health and Psychosocial Support;
FGDs: Focus Group Discussions;
KIIs: Key Informant Interviews;
LMS: Learning Management System;
FRAME: Framework for Reporting Adaptations and Modifications-Enhanced;
LHWs: Lady Health Workers

## Footnotes

1 (Humayun et al., 2025).

2 (World Health Organization, 2018)

3 World Health Organization. (2018). *Problem management plus (PM+): individual psychological help for adults impaired by distress in communities exposed to adversity. (Generic field-trial version 1.1)*. https://www.who.int/publications/i/item/problem-management-plus-(-pm-)-individual-psychological-help-for-adults-impaired-by-distress-in-communities-exposed-to-adversity

4 Humayun et al., 2020. https://journals.lww.com/invn/fulltext/2020/18020/designing_psychosocial_support_for_covid_19.7.aspx

5 (Goldberg & Williams, 1988). https://www.scirp.org/reference/ReferencesPapers?ReferenceID=189650

6 During the feasibility testing in ICT, the course completion report showed that some participants completed the course over more than one session (taking more than 2 hours.

