## Supplementary materials for "Hamdard Force: A scalable, data-informed community-based intervention to develop a mental health workforce in Pakistan"

**Supplementary Material**

**A- Adaptation semi-structured questionnaire (ADAPT checklist)**

a. What adaptations can be proposed in response to identified constraints and facilitators?
b. What cultural considerations are important in adapting PFA guide for our community setting?
c. Which cultural beliefs, idioms of distress, or social norms, rights of people need to be considered during adaptation?
d. How can the PFA Guide be more accessible, and feasible, considering different levels of literacy and learning preferences in our context?
e. What resources are needed to deliver an adapted intervention?
f. Which scalable modalities can be used to train a workforce in resource-limited settings while maintaining quality?
g. How do we measure training efficacy?
h. What adaptations are needed for the sustainability of the intervention?

**B- Inclusion criterion for recruiting Hamdard Force:**

1. High school education (FSc /A Levels / intermediate)
2. Active engagement with the community
3. Effective communication skills
4. Commitment to promote mental health in the community
5. Aspiration for continuous professional development
6. Good standing in their communities

**C- FRAME Reporting**

| **FRAME reporting adaptations** | | |
| --- | --- | --- |
| 1 | When the modification was made | Pre-implementation/planning/pilot |
| 2 | Whether the modification was planned/proactive or unplanned/reactive, | Planned/proactive |
| 3 | Who participated in the decision to modify?  Optional: Who made the ultimate decision | The modifications were planned by the core team based on FGDs/KIIs. Part of the team included community members, recipients, health administrator, intervention team, practitioners.  Experts team finalized the modifications. |
| 4 | What is modified?  Type or nature of context or content-level modifications | Content- addition, deletion, tailoring, shortening, substituting  Structural- format, reordering, spreading  Training an evaluation – format, setting, adding elements Implementation and scale-up activities – delivery mode, mobile application development, system level integration (see tables) |
| 5 | At what level of delivery the modification is made | Target intervention group – community level |
| 6 | The extent to which the modification is fidelity-consistent | Core elements of original content is preserved |
| 7 | The reasons for the modification, including  (a) the intent or goal of the modification (e.g., improve fit, adapt to a different culture, reduce costs, etc.) and  (b) contextual factors that influenced the decision. | Rationale is presented in Table X with each modification.  it broadly covers modifications due to improve fit to a new culture and setting. |

**D. Consort diagram of recruited Hamdard Force**

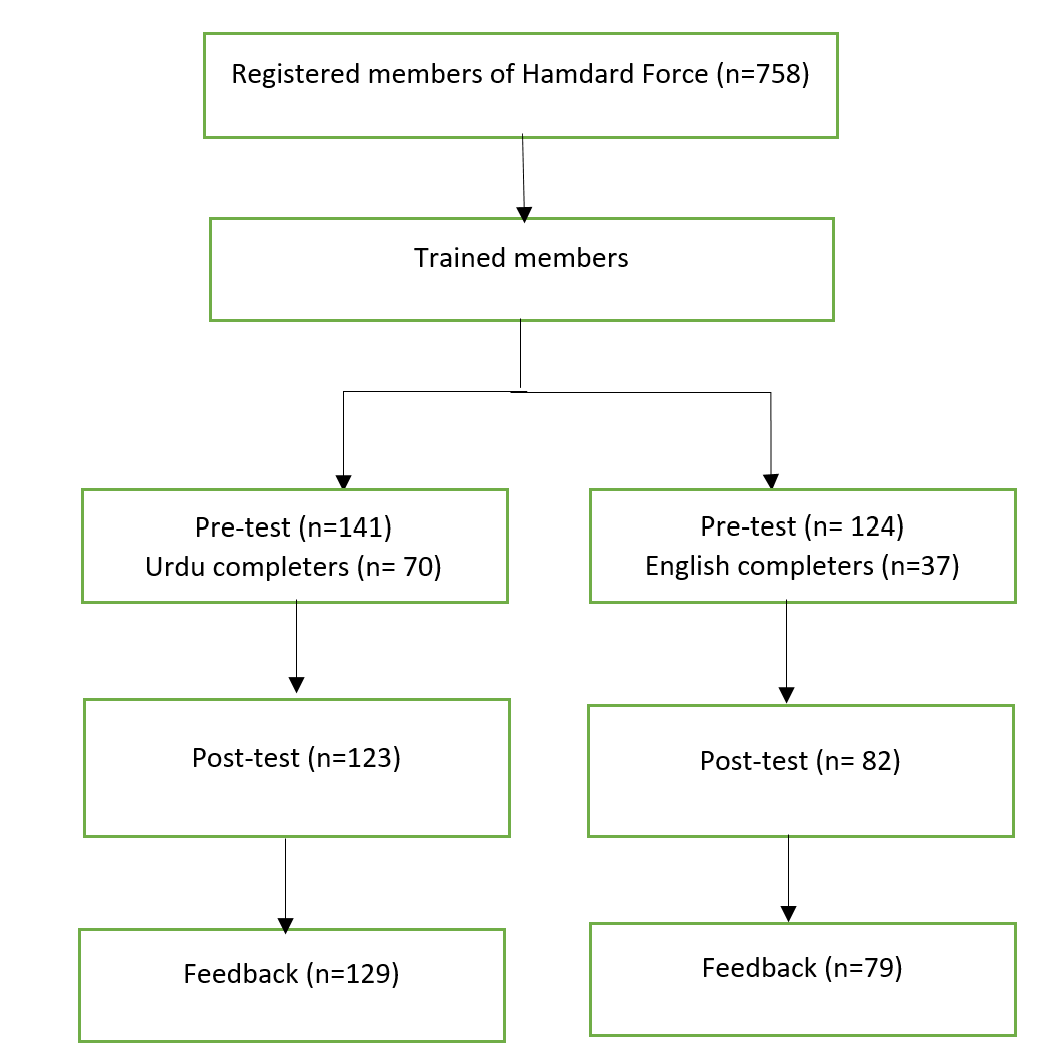
